# Anti-PF4 levels of patients with VITT do not reduce 4 months following AZD1222 vaccination

**DOI:** 10.1101/2021.08.17.21262138

**Authors:** Phillip L.R. Nicolson, Samantha J. Montague, Christopher W. Smith, Clare S. Lodwick, Charlotte Stoneley, Matthew Roberts, Steve P. Watson, Gillian C. Lowe, William A. Lester

## Abstract

**Background:** Anti-Platelet Factor 4 (PF4) IgG antibodies that activate platelets via FcγRIIa have been shown to be an important part of the pathophysiology of vaccine-induced immune thrombocytopenia and thrombosis (VITT). There is now extensive literature on its presentation and initial management. There is no literature however on what happens to these patients following discharge.

**Methods:** We collected clinical data and samples from seven patients presenting with VITT and followed them up for 82-145 days. We also collected clinical samples from them at last follow-up. Testing for anti-PF4/heparin antibodies was done using an anti-PF4/heparin enzymatic immunoassay. Flow Cytometry was used to look at FcγRIIa levels on patient platelets. Light Transmission Aggregometry with patient serum and healthy donor / patient platelets was used to analyse platelet responsiveness, in the presence and absence of PF4.

**Findings:** All patients were discharged on direct oral anticoagulants. Two patients remain completely symptom free, three have ongoing headaches, two have residual neurological deficits. Two patients developed mild thrombocytopenia and worsening headache (but without cerebral venous sinus thrombosis) and were retreated, one of these with rituximab. All patients, except the one treated with rituximab, had similar anti-PF4 antibody titres at 80-120 days to their levels at diagnosis. Platelets from patients at follow-up had normal levels of FcγRIIa and had normal responses to thrombin and collagen-related-peptide. Patient serum from diagnosis strongly activated healthy donor platelets in the presence of PF4. Serum from follow-up was much weaker at stimulating platelets, even in the presence of PF4.

**Interpretation:** This study shows that despite similar PF4 antibody titres at diagnosis and during follow-up, there are further differences in patient serum, that are not apparent from currently used testing, that result in lower levels of platelet activation during the follow-up period. Further understanding of these factors are important in order to assess duration of anticoagulation for these patients.

**Funding:** This work was supported by an Accelerator Grant (AA/18/2/34218) from the British Heart Foundation (BHF) and by a National Institute for Health Research (NIHR) grant.

**Key points:**

1. PF4 antibody titres do not reduce up to 4-months post ChAdOx1 nCoV-19 in patients with VITT
2. Despite similar PF4 antibody titres, diagnostic serum is more potent at activating platelets in the presence of PF4 than follow-up serum.

## Introduction

SARS-CoV-2 was declared a global pandemic in March 2020 (1). A major part of the fight against the pandemic is vaccination. These comprise mRNA vaccines such as those developed by Pfizer-BioNTec and adenoviral vector vaccines such as ChAdOx1 nCoV-19 (AZD1222) which has been developed by Oxford-AstraZeneca (2,3). These vaccines began rolling out worldwide in late 2020 to early 2021. A new syndrome of vaccine-induced thrombocytopenia and thrombosis (VITT), which had a striking similarity to spontaneous Heparin Induced Thrombocytopenia (HIT) was first described in April 2021 (4,5,6,7). The majority of cases occur 4-30 days following AZD1222 with a minority of cases described following the Johnson and Johnson vaccine Ad26.COV2.S (8). They are characterised by thrombocytopenia, high D-dimer and aggressive thrombosis with the majority of clinically significant clots forming at unusual venous sites such as within the cerebral venous sinuses and splanchnic veins but arterial thromboses have been described (9,10). Like patients with HIT, patients with VITT have also been found to have very high titres of antibody to Platelet Factor 4 (PF4) (7). However, unlike with HIT antibodies, these antibodies bind to a single site on PF4 (11). Serum from these patients has been shown by us and other groups to activate platelets via the low affinity FcγRIIa receptor (5,10,12).

Adenoviral vector vaccines are a key part of the worldwide vaccination strategy because they have a relatively low cost and do not have the challenging storage requirements of the mRNA-based vaccines. Understanding VITT, therefore, is of critical importance in order to enable successful mass vaccination, particularly in the developing world.

New treatment modalities for VITT have evolved rapidly and now include non-heparin anticoagulation, corticosteroids, intravenous immunoglobulin (IVIg) and thrombectomy (13). Plasma Exchange (PEX) and the anti-CD20 monoclonal antibody rituximab have been used in patients who fail to respond to first line therapies (9).

Despite treatment, mortality remains in the region of 20-50% (14,15) but those that do respond tend to do so quickly, and are discharged on non-heparin anticoagulation (13). At present the natural history of VITT is unknown. Many questions about its medium to long term prognosis and treatment remain unanswered. Here we describe the clinical course, laboratory results and anti-PF4 antibody levels of 7 patients with VITT who survived their initial presentation and have been followed up for more than 3 months following their triggering vaccination. We also report changes over time in the ability of serum from these patients to activate their own and healthy donor platelets *in vitro*.

### Case series

The presenting clinical features and laboratory results of 5 of the 7 patients below have been previously reported (12). A brief summary is included in Table 1.

**Table 1.** Summary of Clinical Characteristics of Patients with VITT.

|  | Patient 1 | Patient 2 | Patient 3 | Patient 4 | Patient 5 | Patient 6 | Patient 7 |
| --- | --- | --- | --- | --- | --- | --- | --- |
| **Age** | 45-49 | 20-24 | 45-49 | 40-44 | 40-44 | 45-49 | 45-49 |
| **Sex** | Male | Male | Female | Female | Male | Female | Female |
| **Platelet count**<br>**at presentation (x10<sup>9</sup>/L)**<br>normal range 150 - 450 | 16 | 113 | 7 | 11 | 35 | 78 | 66 |
| **D-dimer**<br>**at presentation (ng/mL)**<br>normal range 0 - 250 | 62342 | 22903 | 31301 | 30324 | 6807 | 14274 | 6035 |
| **Fibrinogen**<br>**at presentation (g/L)**<br>normal range 1.5 - 4 | 1.2 | 0.98 | 1.1 | 1.07 | <0.35 | 1.54 | 4.09 |
| **Anti-PF4 antibody titre**<br>**at presentation**<br>**(Optical density)**<br>normal range 0.01 - 0.4 | 2.45 | 2.80 | >3.0 | 1.77 | 2.58 | >3.0 | >3.0 |
| **Site of thrombosis** | CVST | Ischaemic stroke | CVST | CVST | CVST | CVST | CVST |
| **Number of days post vaccine**<br>**at presentation** | 14 | 10 | 14 | 11 | 9 | 20 | 20 |

CVST, cerebral venous sinus thrombosis.

Patient 1 (who suffered a CVST) was discharged on dabigatran. At this point he had a mild headache and some ongoing pyramidal weakness which has gradually resolved over the subsequent three months. His D-dimer and fibrinogen have stabilised in the normal range. In response to a persistent mild thrombocytopenia and increasing headaches (without recurrence of CVST) he was retreated with 1 g/kg IVIg. After this his platelet count increased to normal and his symptoms ceased. His high PF4 antibody level at diagnosis has not significantly changed at >100 days post vaccination (see Figure 1A).

**Figure 1.**
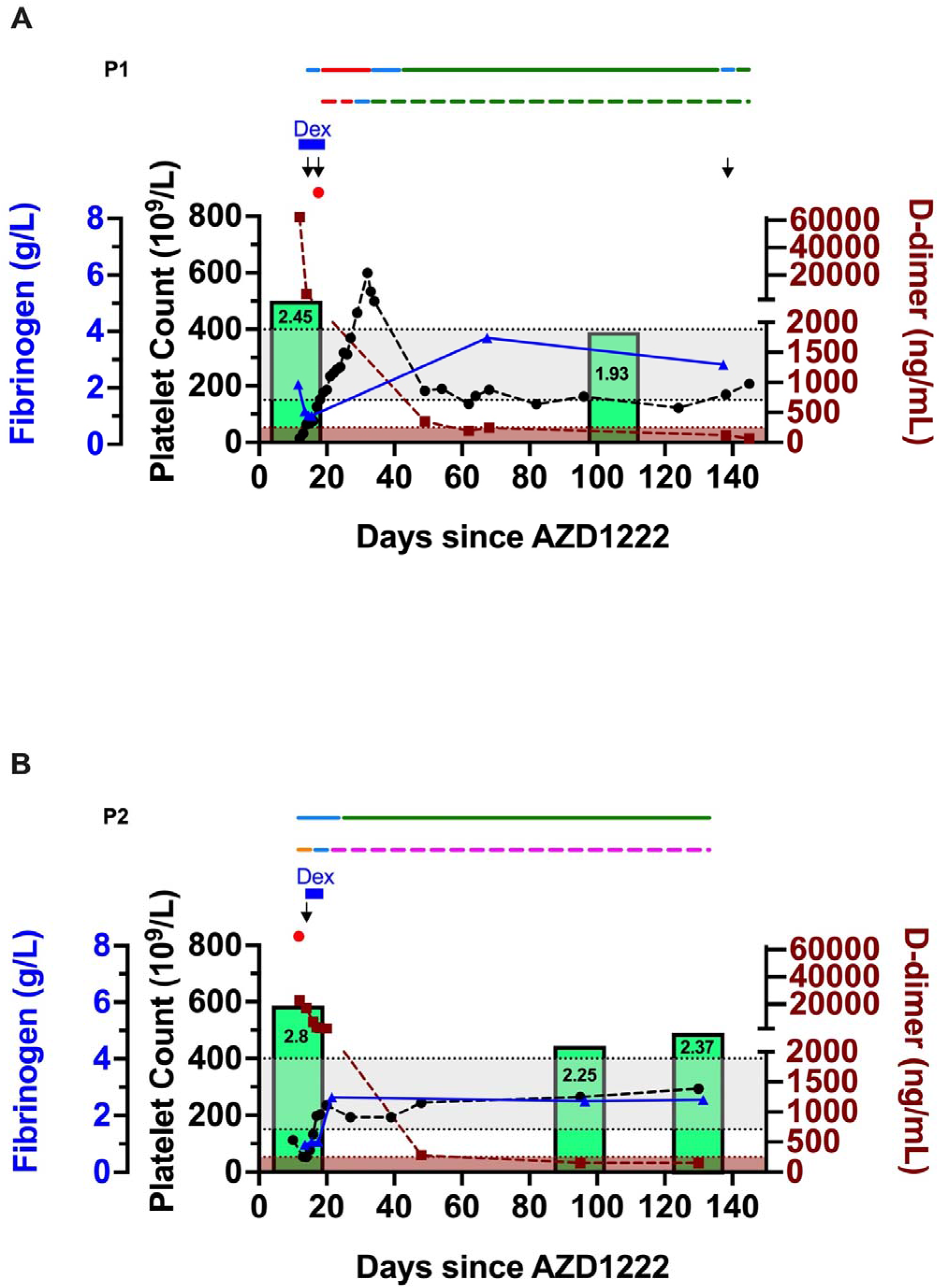
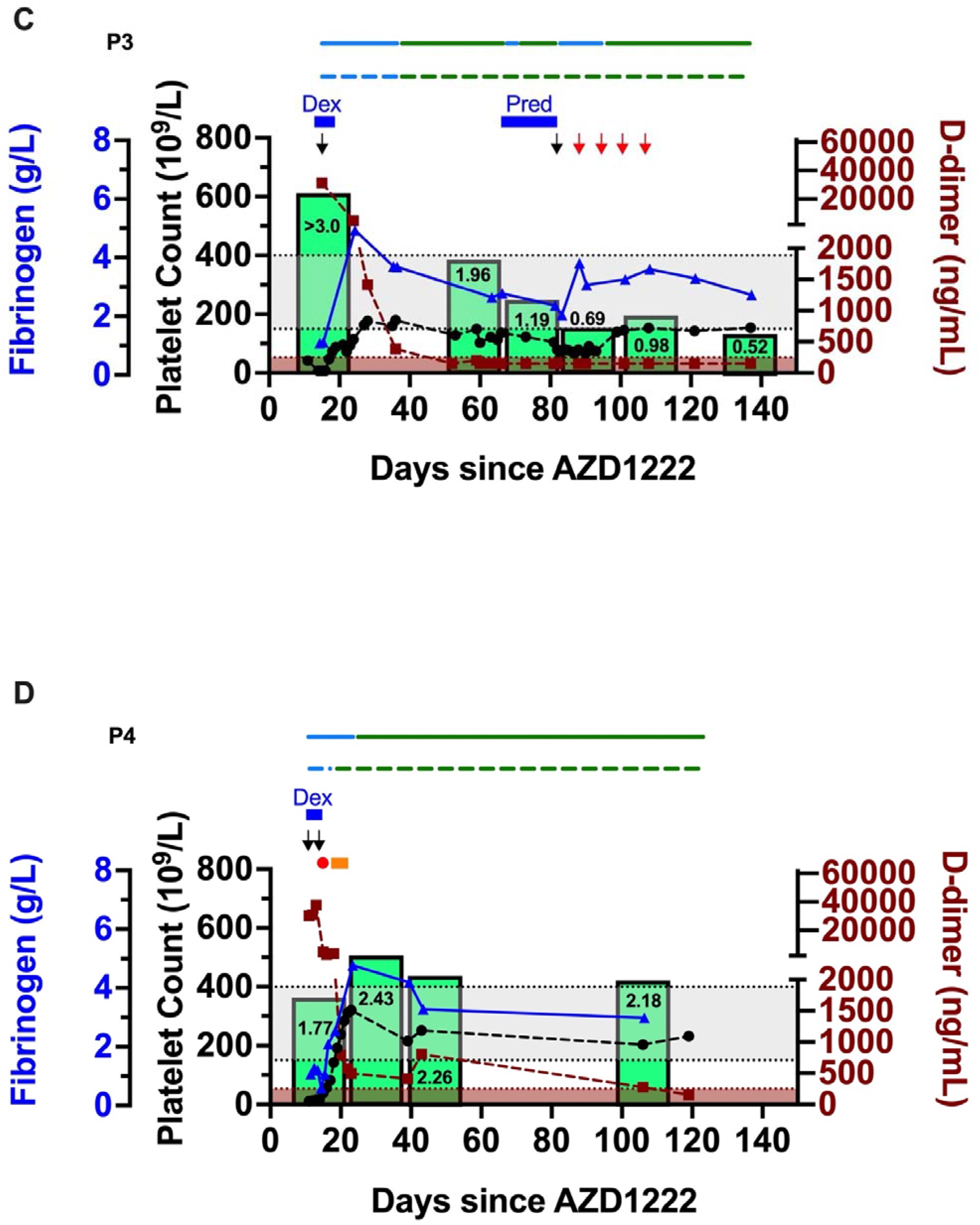
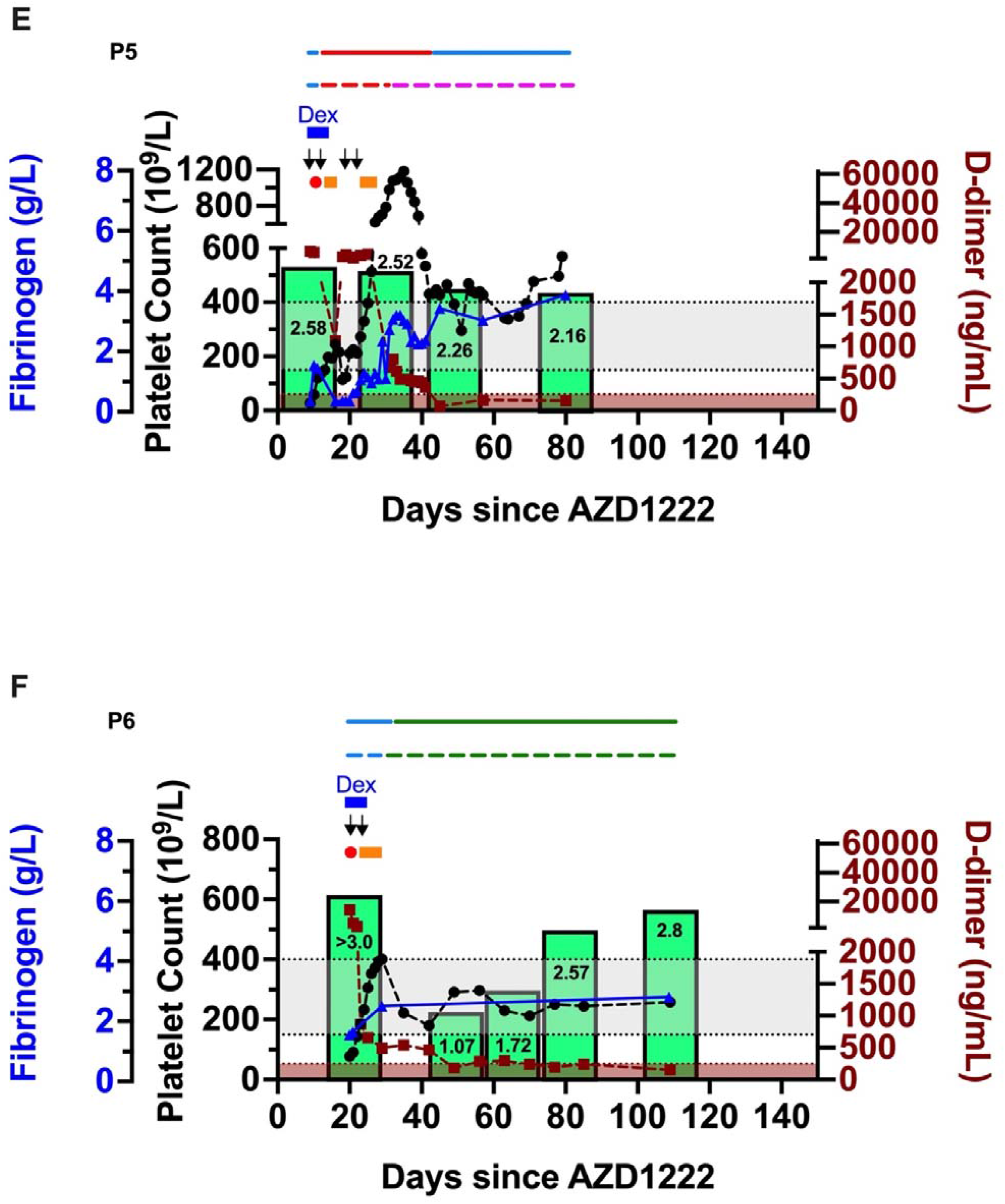
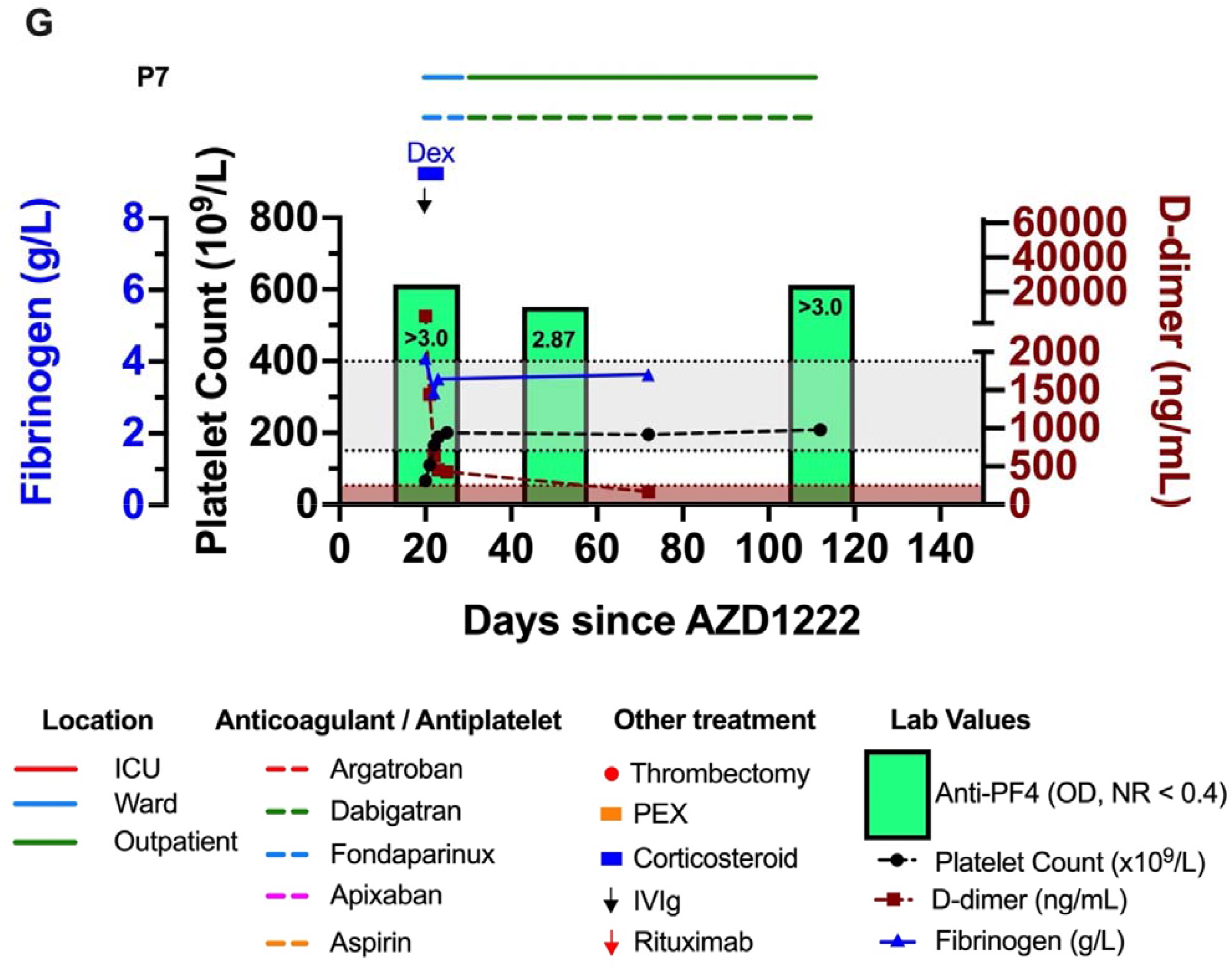
Laboratory parameters of patients with VITT up to 145 days following vaccination with AZD1222. Grey box denotes normal range for platelet count and fibrinogen, brown box denotes normal range for D-dimer. Dex, Dexamethasone. Pred, Prednisolone. ICU, Intensive Care Unit. PEX, Plasma Exchange. IVIg, Intravenous immunoglobulin. PF4, Platelet Factor 4. OD, optical density. NR, normal range.

Patient 2 suffered a middle cerebral artery (MCA) infarct. Following discharge on apixaban he has had ongoing mild unilateral weakness. This is gradually resolving. He did not have a headache. His platelet count, D-dimer and fibrinogen have remained normal following discharge. His PF4 antibody level has remained >2.0 OD throughout (see Figure 1B).

Patient 3 was discharged on dabigatran after her CVST. She had ongoing problems with headaches requiring readmissions. These admissions were associated with recurrence of mild thrombocytopenia but a normal D-dimer and fibrinogen. She was treated with prednisolone and subsequently IVIg and rituximab. Following these further treatments her platelet count normalised, her headache resolved and her PF4 antibody titre dropped to 0.5 OD (see Figure 1C).

Patient 4 suffered a CVST and underwent thrombectomy, IVIg and plasma exchange before being discharged on dabigatran. Now ∼120 days following discharge she is completely symptom free, has a normal platelet count, D-dimer and fibrinogen. Her follow-up PF4 antibody titre has increased since her diagnosis and remains >2.0 OD (see Figure 1D).

Patient 5 was discharged for specialist neurological rehabilitation after a long admission following his initial presentation with CVST. He relapsed early his initial therapy and required re-treatment. He then developed infection with resultant thrombocytosis. Following successful treatment of his infection his platelet count, D-dimer and fibrinogen have been normal but his anti-PF4 antibody titres remain persistently >2.0 OD (see Figure 1E).

A sixth patient (a female aged 45-49) who was not included in our previous study developed VITT and suffered a CVST 20 days following her first vaccination with AZD1222. Her presenting PF4 antibody titre was >3.0 OD. She was initially treated with fondaparinux, dexamethasone and IVIg but also required thrombectomy and plasma exchange. She was discharged on dabigatran. Despite an initial drop in her PF4 antibody titre following discharge, this has risen again and, at >100 days following vaccination, has reached similar levels to those at her presentation but her platelet count and D-dimer remain normal (see Figure 1F). She has an ongoing mild headache.

A seventh patient (a female aged 45-49) who was also not included in our previous study, presented with VITT and resultant CVST 20 days following AZD1222. She was treated with fondaparinux, dexamethasone and IVIg and was discharged on dabigatran. Her platelet count, D-dimer and fibrinogen level have remained normal and at latest follow-up (∼110 days following vaccination) she had no symptoms but still had a PF4 antibody titre of >3.0 OD (see Figure 1G).

## Methods

### Patients and ethical approval

Patients presenting with thrombosis and thrombocytopenia, occurring after AZD1222 vaccination were recruited. Informed consent was provided by the patients or by their next of kin in those who lacked capacity. Collection of blood from these patients (including those lacking capacity) was approved under research ethics granted to the University of Birmingham Human Biomaterial Resource Centre (North West – Haydock Research Ethics Committee, reference: 15/NW/0079, amendment 3, 19/11/2018). Ethical approval for collecting blood from healthy volunteers was granted by Birmingham University Internal Ethical Review Committee (reference: ERN_11-0175, assessed 22/3/2021). All studies were performed in line with the Declaration of Helsinki.

### Materials

PF4 was from Chromatec GmbH (Greifswald, Germany), collagen-related-peptide (CRP) was from CambCol Ltd (Ely, UK), FITC-conjugated mouse anti-human CD32a antibody was from BD Pharminogen (Wokingham, UK), FITC-conjugated IgG1 control mouse antibody was from Dako (Santa Clara, CA). All other reagents were from Sigma-Aldrich.

### Serum preparation

Patient and healthy donor serum was collected following centrifugation (2000 *g*, 10 minutes, room temperature [RT]) of clotted whole blood.

### PF4 antibody testing

Testing for anti-PF4/heparin antibodies was done using an anti-PF4/heparin enzymatic immunoassay (EIA, LIFECODES PF4 enhanced assay; Immucor GTI Diagnostics) for IgG, IgM, and IgA PF4-heparin antibodies.

### Platelet preparation

Preparation of washed platelets from citrated whole blood has already been described. (16) Briefly; citrated blood was taken from healthy, drug-free volunteers and mixed (10:1, v/v) with acid citrate dextrose and centrifuged (200 *g*, 20 minutes, RT) to produce platelet rich plasma. Platelet rich plasma was then centrifuged (1000 *g*, 10 minutes, RT) in the presence of 0.2 μg/mL prostacyclin. The platelet pellet was resuspended in modified-Tyrode’s-HEPES buffer, acid citrate dextrose and 0.2 μg/mL prostacyclin and centrifuged (1000 *g*, 10 minutes, RT). Platelet pellet was resuspended in modified-Tyrode’s-HEPES buffer to a concentration of 2×10^8^/mL and allowed to rest for 30 minutes prior to use.

### Light transmission aggregometry (LTA)

Aggregation was measured in washed platelets (2×10^8^/mL) under stirring conditions (1200 rpm) at 37°C using a light transmission aggregometer (Model 700, ChronoLog) for 30 minutes following stimulation with serum (14:1, v/v) in the presence or absence of 10 μg/mL PF4.

### Platelet FcγRIIa surface measurements

Platelet FcγRIIa (CD32a) surface measurements were obtained using flow cytometry (BD Accuri C6). Washed platelets (2×10^7^) were stained for 30 min with mouse anti-human CD32a antibody or FITC control antibody.

### Statistical analysis

All data are presented as mean ± standard deviation (SD).

## Results

We and others have previously shown that serum from patients with VITT with high anti-PF4 titres activates healthy donor washed platelets (5,12). PF4 antibody titres remained high in all 7 patients (but reduced in one patient treated with rituximab) but this was not associated with clinical evidence of relapse (see Figure 1). Studies were undertaken to investigate whether there was a change in the responsiveness in the patient’s own platelets over time that might make them less able to be activated. We isolated washed platelets from patients 4, 6 and 7 at their most recent follow-up (Day 119, 109 and 112 post AZD1222 respectively). Given platelet activation in patients with VITT has been shown to be mediated via FcγRIIa we used flow cytometry to look at the levels of this receptor on patient platelets and compared this with healthy controls. There was no difference between the median fluorescence intensity on patients or healthy controls (see Figure 2A). We also tested the patient platelets to see if they would respond to G protein-coupled receptor (GPCR) and tyrosine kinase-linked receptor (TKR) agonists thrombin and CRP. Their responses were normal compared to healthy controls (see Figure 2A).

We then investigated whether the follow-up patient serum still had the ability to activate platelets. We took follow-up serum at the same time as PF4 antibody sampling was performed in all 7 of our patients, i.e 80-138 days following AZD1222 vaccination. We then compared levels of healthy donor platelet activation by diagnostic and follow-up serum for each patient in the presence or absence of 10 μg/mL PF4 by LTA. Platelet activation in response to diagnostic time point serum was stronger than that caused by follow-up time point serum (see Table 2). PF4 caused marked potentiation of platelet activation to the diagnostic time point serum but either caused no or low-level potentiation to follow-up serum (see Figure 2B).

**Table 2.**
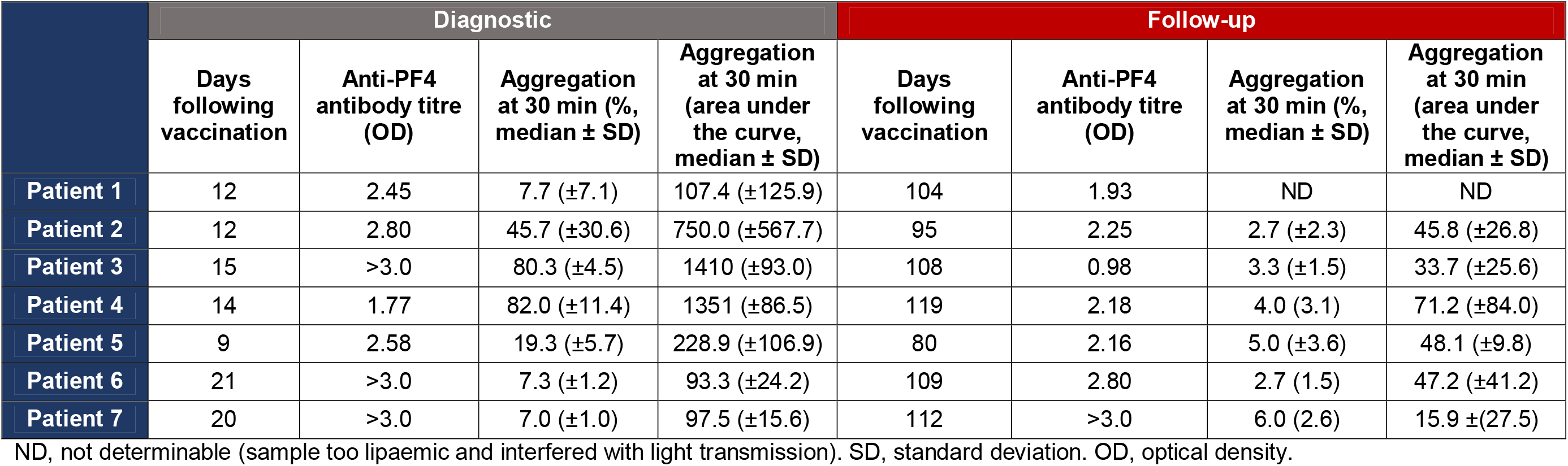
Patient serum activates washed platelets more strongly at diagnosis than it does at follow-up. Washed platelets (2×10^8^/mL) were isolated from healthy donors and warmed for 5 minutes at 37 °C before being stimulated with undiluted patient serum (ratio 14:1) and aggregation measured by light transmission aggregometry for 30 minutes. n=3 for all experiments.

We then looked at patient platelet responses in patients 4, 6 and 7 to their own serum in the presence or absence of PF4, both from diagnosis and from the most recent follow-up time point. Like the results in healthy donor washed platelets, we found that the patient’s own diagnostic serum either caused robust aggregation on its own (patient 4), or did so in the presence of 10 μg/mL PF4 (patients 6 and 7) but that their follow-up serum caused no platelet activation on its own, and that this was not potentiated by PF4 in patient 4, and only subtly potentiated in patients 2 and 7 (see Figure 2C).

## Discussion

This study describes the clinical features and laboratory parameters of patients with VITT in the months following their acute presentation. We show that despite immunosuppressive treatment with corticosteroids the anti-PF4 antibody titres remain strongly positive up to 4 months following AZD1222 vaccination. Anti-PF4 antibody levels in patients with HIT(T) are reported to persist up to 150 days. (4) Only the patient treated with rituximab displayed a reduction in PF4 antibody levels. All except two patients remained in remission with normal platelet counts and very little by way of symptoms despite this high antibody level. The two patients that relapsed only developed mild thrombocytopenia and headache but did not have recurrence of CVST. D-dimers and fibrinogen levels were also normal which is explained by the ongoing use of anticoagulation.

The serum from the follow-up samples of these patients did not activate platelets. This result cannot be easily explained. The patient’s platelets remained strongly reactive to GPCR and TKR agonists, to their own diagnostic serum and had normal FcγRIIa levels. These results imply that a high PF4-antibody titre alone is not sufficient to cause VITT and that an additional feature present in the patient serum at diagnosis, but not at follow-up, is required for VITT to develop. This potentially could be explained by reduction in PF4 levels in patients over time, by the ongoing use of anticoagulation or by the consumption of high-affinity PF4 antibodies. The observation that serum samples can still be made to activate platelets in the presence of PF4 raises concern that any inflammatory or traumatic event that results in the release of PF4 may cause a relapse of thrombocytopenia. It may therefore be necessary to continue anticoagulation for as long as there are raised PF4 antibody titres. These results also indicate that rituximab may be an appropriate treatment for corticosteroid and IVIg resistant relapse. This area requires further study before any recommendations about the medium- to long-term treatment of these patients can be made.

## Data Availability

Individual participant data will not be made available.

## Acknowledgements

This work was supported by an Accelerator Grant (AA/18/2/34218) from the British Heart Foundation (BHF) and a National Institute for Health Research (NIHR) grant. SPW holds a BHF Chair (CH03/003).

## Authorship

PLRN recruited patients, collected and analysed clinical data, designed and performed experiments and wrote the manuscript. SJM and CWS designed and performed experiments and analysed data. CSL collected clinical data. CS and MR performed experiments. SPW designed experiments. GCL and WAL recruited patients and contributed intellectually. All authors revised the manuscript. Underlying data was verified by PLRN, SJM and CWS.

---

**Supplementary Figure 1:**
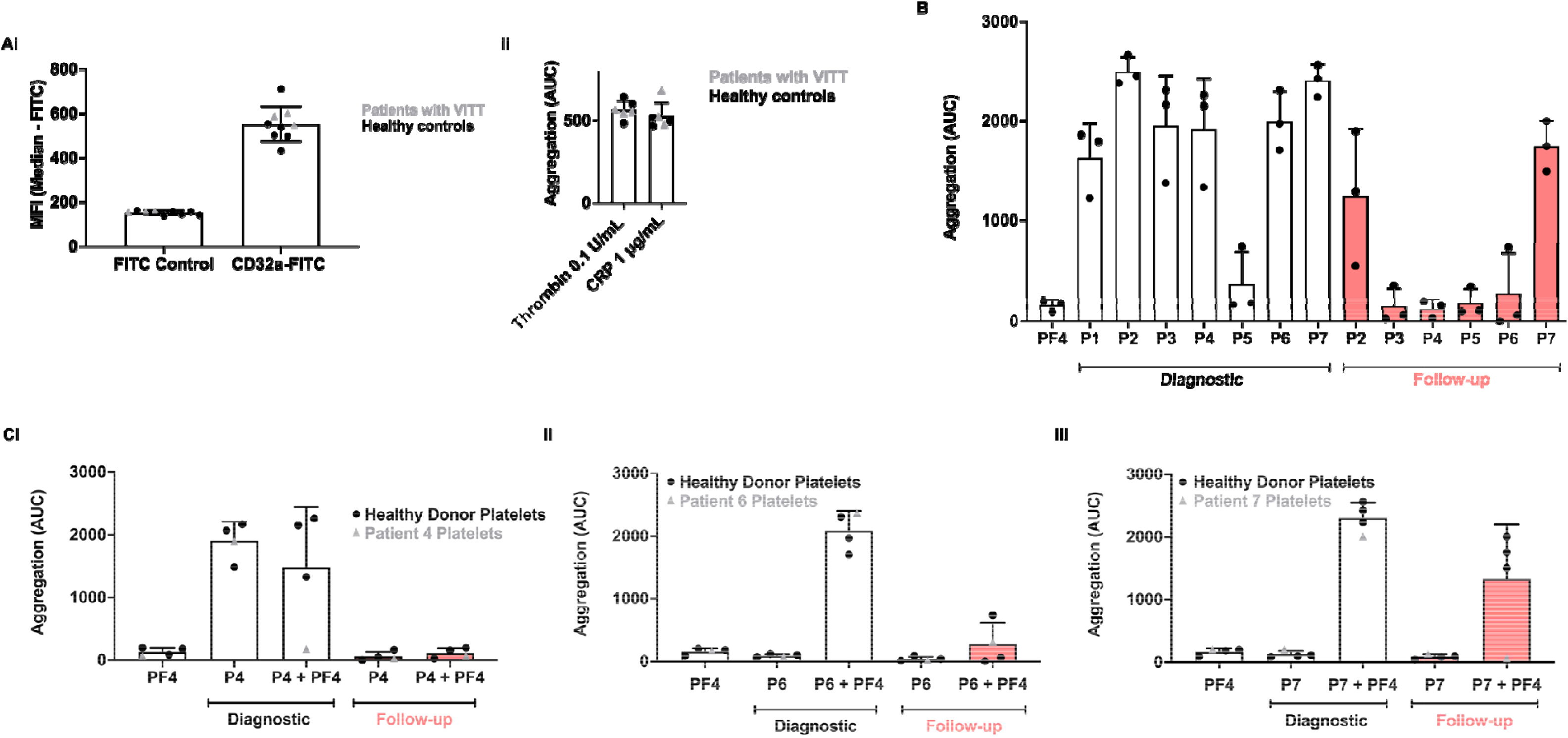
PF4 potentiation of VITT patient sera-induced aggregation is greater with diagnostic samples compared to follow-up samples. Washed platelets (2 ×10^8^/mL) were isolated from patients with VITT 109 – 119 days following vaccination or from healthy donors. **Ai** Conjugated a CD32a (FcγRIIa) antibody was added to patient (triangle) and healthy donor (circle) washed platelets for 30 minutes before termination with excess phospha buffered saline and median fluorescence intensity measured. N=5 healthy controls and n=3 VITT patients (P4, P6, P7), median ± SD. **ii** Washed platelets (2 ×10^8^/mL) from patients and healthy donors were incubated at 37 °C for 5 minutes before being stimulated with thrombin (0.1 U/mL) or collagen related peptid (CRP) (1 μg/mL) under stirring conditions (1200 rpm) for 7 minutes and aggregation measured by LTA. **B** Healthy donor washed platelets were warmed to 3 °C and incubated with PF4 (10 μg/mL) for 5 minutes and then stimulated with patient serum (either taken at diagnosis or taken at most recent follow-up, see Table 2) at 14:1, v:v under stirring conditions (1200 rpm) for 30 minutes. NB P1 not tested at follow-up due to lipaemic serum which interfered with LTA. **C** Healthy donor and patient washed platelets (from patient 4, 6 and 7) were warmed to 37 °C and incubated with PF4 (10 μg/mL) or vehicle (Tyrode’s buffer) f minutes and then stimulated with diagnostic and follow-up patient serum (**i** Patient 4, **ii** Patient 6, **iii**, Patient 7). All aggregations are shown as Area Under th Curve as Mean + SD from 3 experiments unless otherwise stated.

